# Parent attitudes and behaviors associated with household food security among rural lower income families

**DOI:** 10.1101/2024.08.22.24312452

**Authors:** Amy M. Moore, Jennifer S. Savage, Melissa N. Poulsen, Jennifer Franceschelli Hosterman, Carolyn F. McCabe, Lisa Bailey-Davis

## Abstract

**Objective:** To examine associations between Theory of Planned Behavior constructs related to parent engagement in health-promoting behaviors and food resource management with household food security status.

**Methods:** A cross-sectional secondary analysis of baseline data from 1622 parents enrolled in the ENCIRCLE study, a pragmatic cluster-randomized controlled trial to prevent obesity among preschool-aged children from rural lower-income families. Logistic regression models examined associations between parent engagement in health-promoting behaviors (attitude, subjective norm, perceived behavioral control) and food resource management (self-confidence, behaviors) with household food security status.

**Results:** Parents with greater perceived behavioral control (OR=1.44, CI:1.14-1.82) and food resource management self-confidence (OR=2.08, CI:1.80-2.41) had significantly higher odds of experiencing household food security in adjusted models.

**Conclusions and Implications:** Findings suggest that perceived behavioral control and food resource management self-confidence may help safeguard lower-income families. Health promotion efforts should target these factors to help parents engage in health-promoting behaviors during times of economic hardship.

## INTRODUCTION

Food insecurity is a complex, dynamic, and persistent social problem. In 2022, 12.8% of US households experienced food insecurity, meaning that these households were “unable to acquire adequate food for one or more household members because they had insufficient money or other resources for food.”^1^ Prevalence of food insecurity is higher among people who identify as Black (22.4%) or Hispanic (20.8%), households with children under 6 years of age (16.7%), headed by a single female (33.1%), located in rural areas (14.7%), and with incomes below the federal poverty level (36.7%).^2^ Experiencing food insecurity during early childhood, a period of rapid growth and development, is associated with negative health outcomes such as poor physical health (e.g., asthma, anemia, and obesity)^3,4^, poor mental health (e.g., depression, negative behaviors, and hyperactivity)^3,5^, and low fruit intake.^6^ Similarly, experiencing food insecurity as an adult is associated with negative health outcomes such as poor physical health (e.g., type 2 diabetes, cardiovascular disease, and obesity)^7–9^, poor mental health (e.g., stress, depression, and anxiety)^10^, and low diet quality.^6^ Parents may also reduce their food intake or quality to protect their children from the effects of food insecurity^11^, potentially worsening these health outcomes. Given that the environmental and socioeconomic determinants of food insecurity are complex and dynamic^12^, not all vulnerable households (e.g., incomes below the federal poverty level) experience food insecurity.^1,2^ Federal nutrition assistance programs can safeguard against food insecurity^13,14^, although less is known about parent attitudes and behaviors that may safeguard against food insecurity during times of economic hardship.

Parents play an important role in shaping their children’s health behaviors. Current public health recommendations include high-quality nutrition, physical activity, and sleep, as well as limited screen time to promote healthy growth and development during childhood.^15,16^ Parents report helping their children establish healthy behaviors as important, although barriers such as cost, limited time, and perceived lack of control may thwart parent engagement in health-promoting behaviors.^17^ In addition, these barriers may be more prevalent in households that experience stressors related to economic hardship and food insecurity. The Theory of Planned Behavior (TPB) is a widely used model to understand health behaviors^18^, including parent engagement in health-promoting behaviors for their young children.^19^ According to the TPB, behavior is determined by the intention to engage in the behavior.^20^ Behavioral intention is determined by three constructs: attitude (i.e., overall positive or negative evaluation of the behavior), subjective norm (i.e., belief that others important to them might approve or disapprove of the behavior), and perceived behavioral control (i.e., belief in the ease or difficulty of performing the behavior).^20^ The more positive the evaluation of these constructs, the stronger the behavioral intention, which in turn increases the likelihood of the behavior. The TPB has been used to understand parent engagement in health-promoting behaviors such as food purchasing^21^, feeding decisions^22^, and obesity prevention^23^ for their young children. However, less is known about parent engagement in health-promoting behaviors to prevent obesity among their young children and household food security status.

Parents also play an important role in shaping the home food environment, including food availability, food purchasing, and food preparation behaviors, which impact child growth and development.^24^ Households experiencing economic hardship face complex and dynamic barriers to sustaining a healthy home food environment, yet some households exhibit resilience and resourcefulness despite these barriers.^25^ Nutrition education and food resource management can help households experiencing economic hardship obtain nutritious and affordable foods.^26,27^ Thus, food resource management behaviors, such as strategies to help families with meal planning, selecting nutritious foods, and stretching limited food budgets, may improve household food and nutrition security.^27^ However, since not all households that experience economic hardship participate in nutrition education programs or experience food insecurity, a more nuanced understanding of the relationship between food resource management and household food security status is needed.

We aimed to identify parent factors that may safeguard households that experience economic hardship from the impacts of food insecurity and promote food and nutrition security. The current study used baseline data from a sample of parents with preschool-aged children living in rural, lower-income contexts to understand parent attitudes, behaviors, and household food security. The first aim used TPB constructs to examine associations between parent engagement in health-promoting behaviors to prevent obesity among their young children and household food security status. The second aim examined associations between parent food resource management self-confidence and behaviors and household food security status. We hypothesized that parents reporting higher scores on TBP constructs, and reporting higher food resource management self-confidence and behaviors, would be associated with household food security.

## METHODS

### Study Design and Sample

This was a cross-sectional secondary analysis of baseline data from the ENCIRCLE study, a pragmatic, cluster-randomized controlled trial designed to test the comparative effectiveness of standard well-child visits versus two enhancements to these visits on obesity prevention among preschool-aged children at risk for obesity living in lower-income households.^28^ The ENCIRCLE study included 2040 parents of preschool-aged children who received care at Geisinger primary care clinics located in predominantly rural areas of central and northeast Pennsylvania that serve socioeconomically diverse patients who are at risk for health disparities. Potentially eligible participants were identified via patient electronic health records and then contacted by the study team for screening. Data were collected for enrolled participants between 2020 to 2023. Children were eligible to participate if they were between 20 to 60 months of age with a BMI-for-age and sex ≥50^th^ percentile and attended annual well child visits at a participating Geisinger clinic. Children were ineligible if they attended annual well-child visits via telemedicine, had a preexisting medical condition (e.g., developmental delays, type 1 diabetes), or had a sibling who participated in the study. Parents were eligible to participate if they were ≥18 years of age, English-speaking, and from households with lower incomes (determined by eligibility for or enrollment in one of the following: Supplemental Nutrition Assistance Program, Special Supplemental Nutrition Program for Women, Infants and Children, Medicaid, Children’s Health Insurance Program, National School Lunch Program, Temporary Assistance for Needy Families, or if they screen positive for household food insecurity). If health insurance was not Medicaid, then this study used an algorithm based on household size, household income, and WIC eligibility criteria during participant screening to confirm lower income status. Details on the study design have been published elsewhere.^28^ Eligible parents provided informed consent to participate. This study was approved by the Institutional Review Board of Geisinger, a large integrated health system, and is registered with ClinicalTrials.gov (NCT04406441).

### Measures

Enrolled parents completed baseline questionnaires using the REDCap electronic data capture tool hosted at Geisinger.^29^ Parents were emailed or texted a REDCap link or were mailed paper questionnaires with a prepaid return envelope upon request. Parents received a $50 gift card upon completing baseline questionnaires.

### Sociodemographic Characteristics

Parents self-reported their sex (female/male), race (American Indian or Alaska Native, Asian, Black or African American, Native Hawaiian or Other Pacific Islander, and White), ethnicity (Hispanic/non-Hispanic), highest level of education (high school diploma or GED, college degree, graduate degree, other), employment (full-time, part-time, unemployed, other), household size (total number in household), and Medicaid enrollment (yes/no; an indicator of family socioeconomic status^30^) at baseline.

### Parent Engagement in Health-promoting Behaviors

Parent engagement in health-promoting behaviors to prevent obesity among young children was assessed at baseline using an adapted version of a measure developed by Andrews et al. that used TPB constructs.^31^ Based on recommendations from the study team and community advisory board members, adaptations were made to the original measure to improve readability for lower literacy audiences. The adapted measure was reviewed by study team members with expertise in nutrition, kinesiology, human behavior, and pediatrics to ensure content validity and community advisory board members to ensure face validity. The adapted 20-item measure assessed parent engagement in health-promoting behaviors to prevent obesity among young children, including limiting sugar-sweetened beverages, sweet snack foods, and screen time and promoting physical activity and sleep across four TPB constructs (attitude, subjective norm, perceived behavioral control, and behavioral intention). Attitude toward health-promoting behaviors was assessed using five items (e.g., “It is important to me to limit my child’s intake of sugar sweetened drinks [such as soda, chocolate milk, or fruit juice] to a small serving or none each day to keep him/her from becoming overweight.”). Subjective norm pertaining to health-promoting behaviors was assessed using five items (e.g., “Most people whose opinions matter to me think that I should limit my child’s intake of sweetened foods to a small serving or none each day.”). Perceived behavioral control of health-promoting behaviors was assessed using five items (e.g., “I am confident that I can limit my child’s screen time to less than 2 hours each day.”). Behavioral intention toward health-promoting behaviors was assessed using five items (e.g., “I intend to get my child to spend his or her free time each day doing something physically active.”). Item response options ranged from 1 (strongly disagree) to 5 (strongly agree). Item responses were averaged across each construct to produce a mean score, with higher scores indicating greater agreement with that construct.

### Food Resource Management

Parent food resource management self-confidence and behaviors were assessed at baseline using 9 items from the SNAP-Ed evaluation framework toolkit.^32^ These items were used and validated in previous studies that assessed the impact of nutrition education programs targeting lower-income audiences on food resource management self-confidence^26,33^ and behaviors.^26,27^ Food resource management self-confidence was assessed using four items (e.g., “How confident are you that you can buy healthy foods for your family on a budget?” and “How confident are you that you can choose the best priced fruits and vegetables?”). Item response options ranged from 1 (not at all confident) to 5 (very confident). Food resource management behaviors was assessed using five items (e.g., “How often do you plan meals ahead of time?” and “How often do you use a grocery list when you go grocery shopping?”). Item response options ranged from 1 (never) to 5 (always). For both subscales, item responses were averaged to produce a mean score, with higher scores indicating greater food resource management self-confidence and behaviors.

### Household Food Security Status

Household food security status during the previous 12 months was assessed at baseline using the 6-item short-form US Household Food Security Survey Module.^34^ The short-form module was selected to reduce participant burden and has acceptable sensitivity and specificity relative to the 18-item module. Scores were calculated by summing affirmative responses for each item resulting in a raw food security score ranging from 0 to 6. Raw scores were categorized as high or marginal food security (0 or 1), low food security (2 to 4), and very low food security (5 or 6). Households were categorized as food secure (included high or marginal food security) and food insecure (included low and very low food security) for analyses.

### Statistical Analysis

Descriptive statistics for the main variables of interest are presented as means and standard deviations for continuous variables and frequencies and percentages for categorical variables. This analysis included 1622 parents who had complete data on household food security at baseline. Independent samples t-tests explored differences in continuous variables and chi-square tests explored differences in categorical variables by household food security status (food secure vs. food insecure). Separate unadjusted logistic regression models examined associations of TPB constructs related to parent intention to engage in health-promoting behaviors to prevent obesity (attitude, subjective norm, perceived behavioral control) with household food security and associations of food resource management self-confidence and behaviors and household food security. For statistically significant unadjusted models, variables that were significantly correlated with household food security status were included in adjusted models (i.e., parent education, P = 0.004 and Medicaid participation, P = 0.009). Data were analyzed using SAS 9.4 and an alpha of P < 0.05 (two-tailed) or a confidence interval not overlapping one were used to determine statistical significance.

## RESULTS

### Sociodemographic Characteristics

Table 1 presents sociodemographic characteristics of study parents by household food security status (n = 1622). Most parents reported sex assigned at birth as female (95.3%), were white (87.7%), had a high school diploma or GED (41.6%), worked full-time or part-time (58.7%), lived in food secure households (82.9%), and participated in Medicaid (65.5%).

**Table 1.**
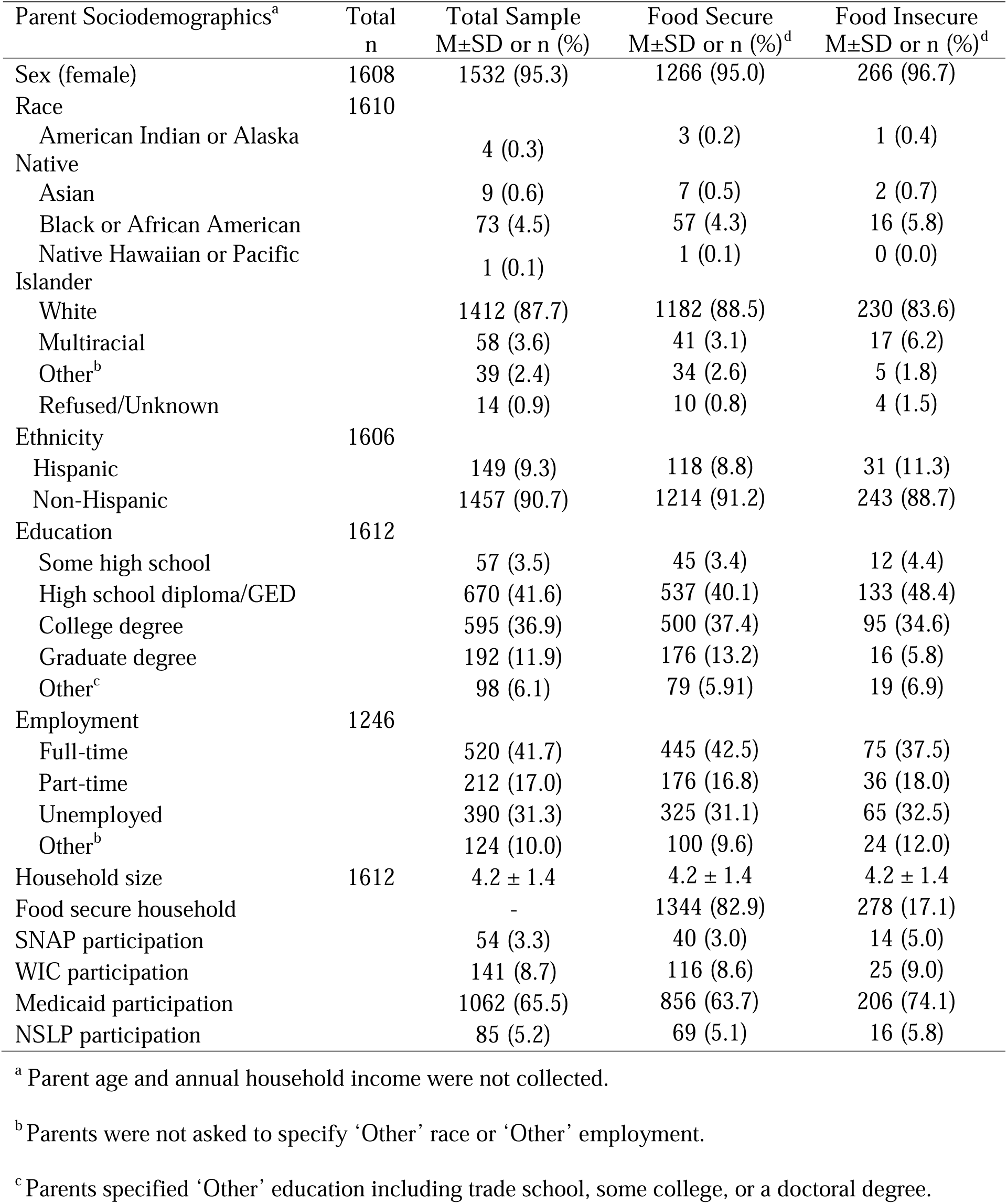
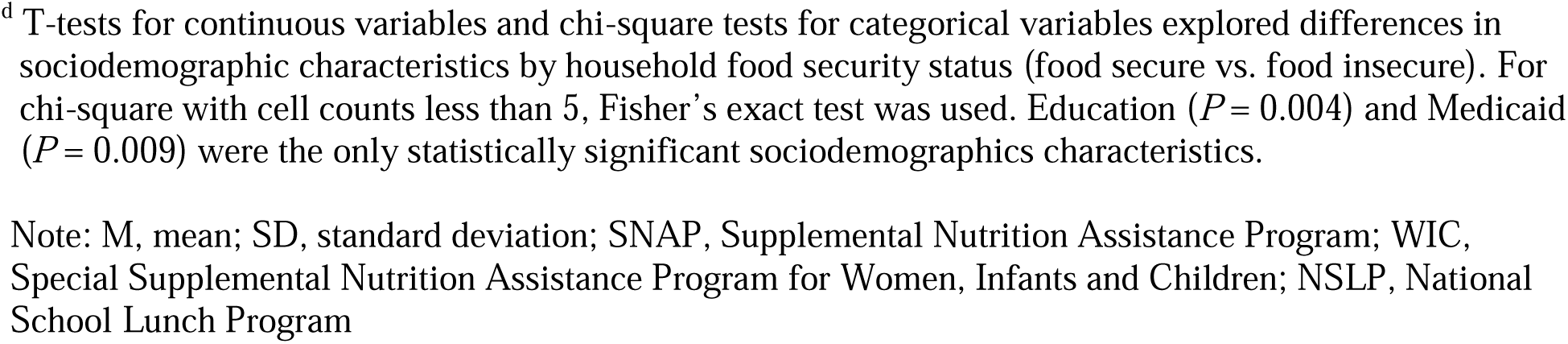
Baseline sociodemographic characteristics for parents in the current study who were enrolled in ENCIRCLE by household food security status (n = 1622).

| Parent Sociodemographics<sup>a</sup> | Total<br>n | Total Sample<br>M±SD or n (%) | Food Secure<br>M±SD or n (%)<sup>d</sup> | Food Insecure<br>M±SD or n (%)<sup>d</sup> |
| --- | --- | --- | --- | --- |
| Sex (female) | 1608 | 1532 (95.3) | 1266 (95.0) | 266 (96.7) |
| Race | 1610 |  |  |  |
| American Indian or Alaska Native |  | 4 (0.3) | 3 (0.2) | 1 (0.4) |
| Asian |  | 9 (0.6) | 7 (0.5) | 2 (0.7) |
| Black or African American |  | 73 (4.5) | 57 (4.3) | 16 (5.8) |
| Native Hawaiian or Pacific Islander |  | 1 (0.1) | 1 (0.1) | 0 (0.0) |
| White |  | 1412 (87.7) | 1182 (88.5) | 230 (83.6) |
| Multiracial |  | 58 (3.6) | 41 (3.1) | 17 (6.2) |
| Other<sup>b</sup> |  | 39 (2.4) | 34 (2.6) | 5 (1.8) |
| Refused/Unknown |  | 14 (0.9) | 10 (0.8) | 4 (1.5) |
| Ethnicity | 1606 |  |  |  |
| Hispanic |  | 149 (9.3) | 118 (8.8) | 31 (11.3) |
| Non-Hispanic |  | 1457 (90.7) | 1214 (91.2) | 243 (88.7) |
| Education | 1612 |  |  |  |
| Some high school |  | 57 (3.5) | 45 (3.4) | 12 (4.4) |
| High school diploma/GED |  | 670 (41.6) | 537 (40.1) | 133 (48.4) |
| College degree |  | 595 (36.9) | 500 (37.4) | 95 (34.6) |
| Graduate degree |  | 192 (11.9) | 176 (13.2) | 16 (5.8) |
| Other<sup>c</sup> |  | 98 (6.1) | 79 (5.91) | 19 (6.9) |
| Employment | 1246 |  |  |  |
| Full-time |  | 520 (41.7) | 445 (42.5) | 75 (37.5) |
| Part-time |  | 212 (17.0) | 176 (16.8) | 36 (18.0) |
| Unemployed |  | 390 (31.3) | 325 (31.1) | 65 (32.5) |
| Other<sup>b</sup> |  | 124 (10.0) | 100 (9.6) | 24 (12.0) |
| Household size | 1612 | 4.2 ± 1.4 | 4.2 ± 1.4 | 4.2 ± 1.4 |
| Food secure household |  | - | 1344 (82.9) | 278 (17.1) |
| SNAP participation |  | 54 (3.3) | 40 (3.0) | 14 (5.0) |
| WIC participation |  | 141 (8.7) | 116 (8.6) | 25 (9.0) |
| Medicaid participation |  | 1062 (65.5) | 856 (63.7) | 206 (74.1) |
| NSLP participation |  | 85 (5.2) | 69 (5.1) | 16 (5.8) |

<sup>a</sup> Parent age and annual household income were not collected.
<sup>b</sup> Parents were not asked to specify ‘Other’ race or ‘Other’ employment.
<sup>c</sup> Parents specified ‘Other’ education including trade school, some college, or a doctoral degree.
<sup>d</sup> T-tests for continuous variables and chi-square tests for categorical variables explored differences in sociodemographic characteristics by household food security status (food secure vs. food insecure). For chi-square with cell counts less than 5, Fisher's exact test was used. Education ( $P = 0.004$ ) and Medicaid ( $P = 0.009$ ) were the only statistically significant sociodemographics characteristics.
Note: M, mean; SD, standard deviation; SNAP, Supplemental Nutrition Assistance Program; WIC, Special Supplemental Nutrition Assistance Program for Women, Infants and Children; NSLP, National School Lunch Program

### Parent Engagement in Health-promoting Behaviors and Food Security

Table 2 presents descriptives for TPB constructs related to parent engagement in health-promoting behaviors to prevent obesity by household food security status. All parents in this study reported high mean values for attitude (M = 4.16, SD = 0.62), perceived behavioral control (M = 4.26, SD = 0.54), and behavioral intention (M = 4.23, SD = 0.56), which were scored on a 5-point scale. The mean value of perceived behavioral control among food secure households (M = 4.28, SD = 0.53) was significantly higher compared to food insecure households (M = 4.17, SD = 0.57; *P* = 0.003). Similarly, the mean value of perceived behavioral intention among food secure households (M = 4.25, SD = 0.56) was significantly higher compared to food insecure households (M = 4.17, SD = 0.57; *P* = 0.02).

**Table 2.**
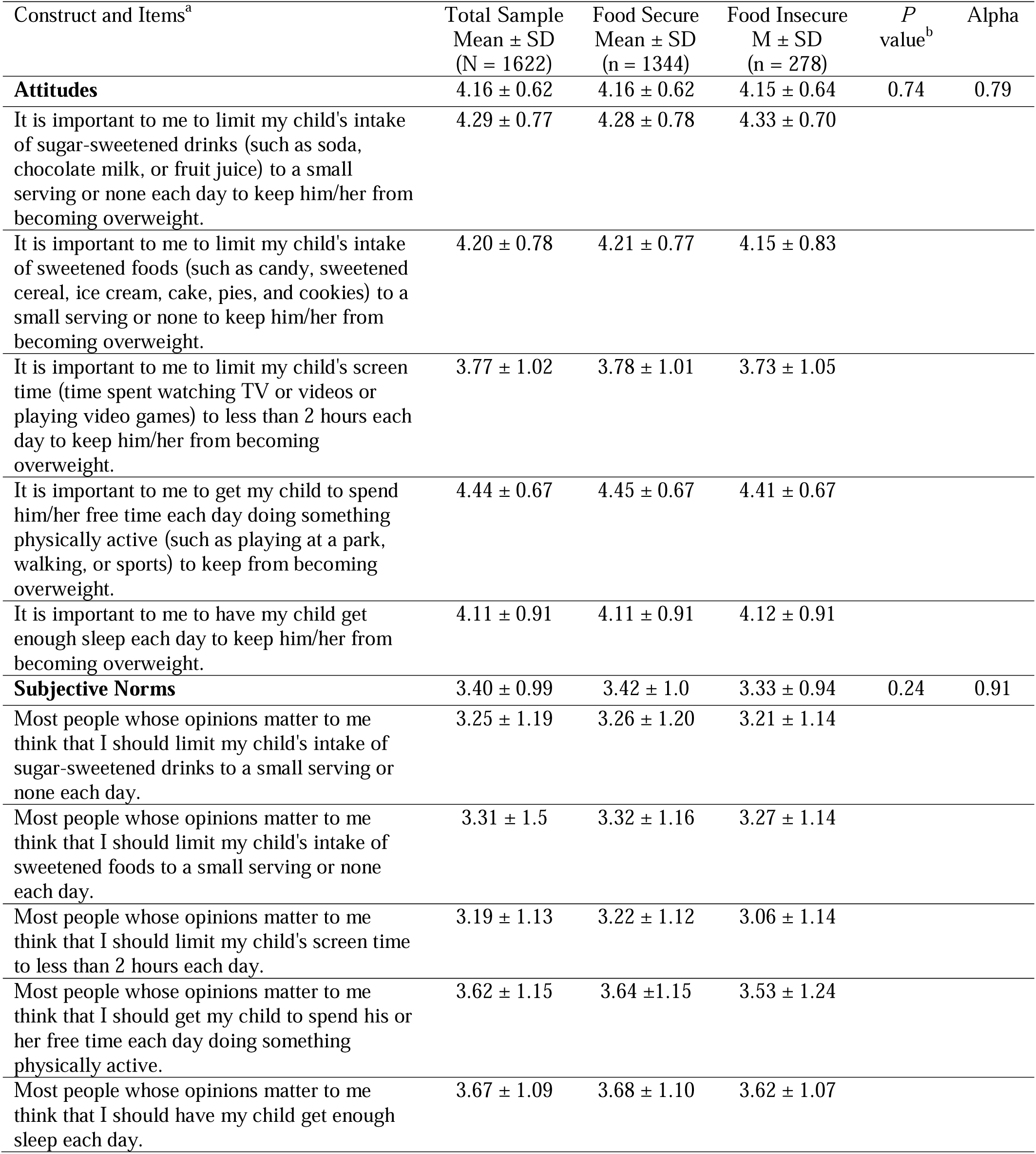
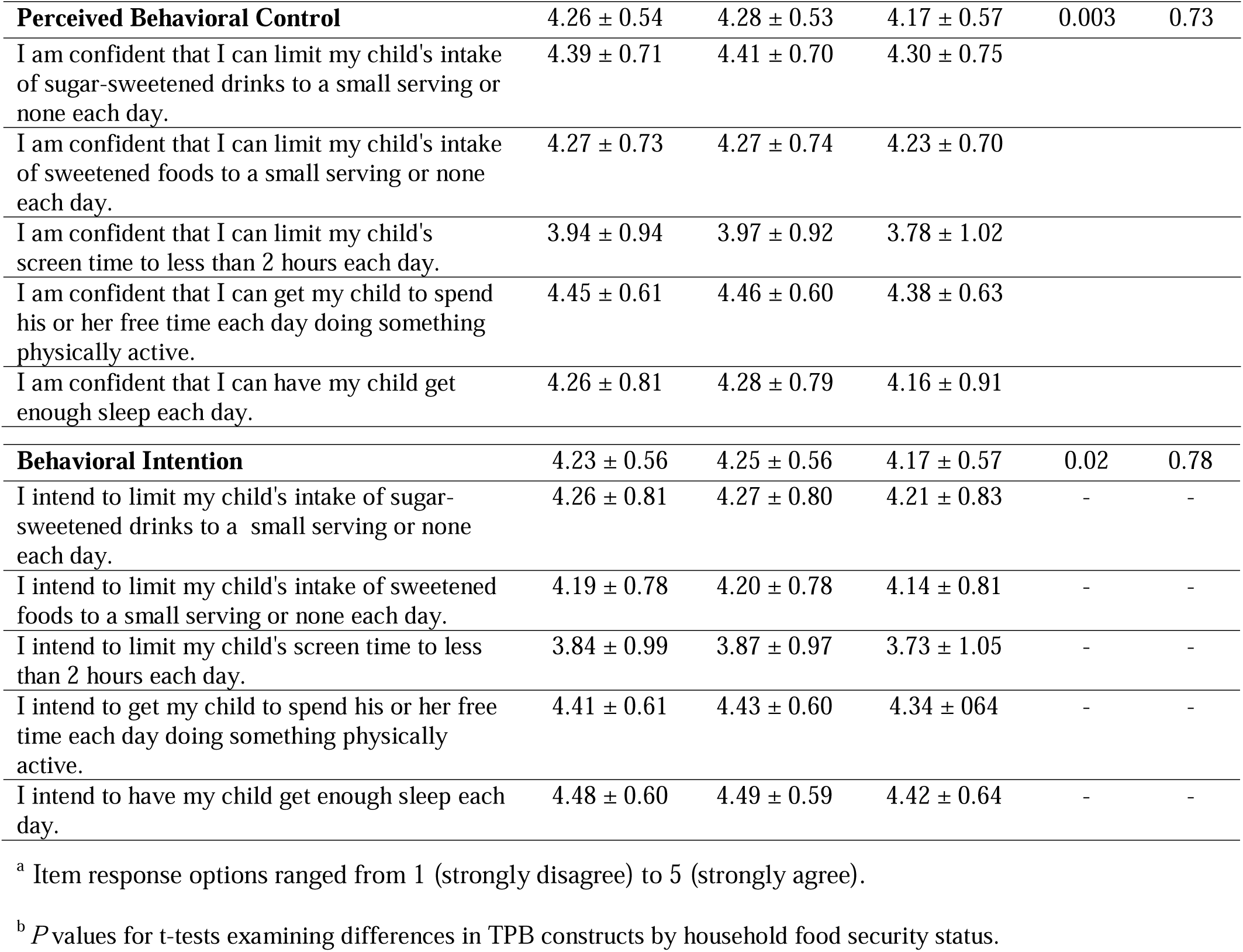
Descriptives for Theory of Planned Behavior (TBP) constructs related to parent engagement in health-promoting behaviors to prevent obesity by household food security status (N = 1622).

| Construct and Items<sup>a</sup> | Total Sample<br>Mean ± SD<br>(N = 1622) | Food Secure<br>Mean ± SD<br>(n = 1344) | Food Insecure<br>M ± SD<br>(n = 278) | P<br>value<sup>b</sup> | Alpha |
| --- | --- | --- | --- | --- | --- |
| **Attitudes** | 4.16 ± 0.62 | 4.16 ± 0.62 | 4.15 ± 0.64 | 0.74 | 0.79 |
| It is important to me to limit my child's intake of sugar-sweetened drinks (such as soda, chocolate milk, or fruit juice) to a small serving or none each day to keep him/her from becoming overweight. | 4.29 ± 0.77 | 4.28 ± 0.78 | 4.33 ± 0.70 | | |
| It is important to me to limit my child's intake of sweetened foods (such as candy, sweetened cereal, ice cream, cake, pies, and cookies) to a small serving or none to keep him/her from becoming overweight. | 4.20 ± 0.78 | 4.21 ± 0.77 | 4.15 ± 0.83 | | |
| It is important to me to limit my child's screen time (time spent watching TV or videos or playing video games) to less than 2 hours each day to keep him/her from becoming overweight. | 3.77 ± 1.02 | 3.78 ± 1.01 | 3.73 ± 1.05 | | |
| It is important to me to get my child to spend him/her free time each day doing something physically active (such as playing at a park, walking, or sports) to keep from becoming overweight. | 4.44 ± 0.67 | 4.45 ± 0.67 | 4.41 ± 0.67 | | |
| It is important to me to have my child get enough sleep each day to keep him/her from becoming overweight. | 4.11 ± 0.91 | 4.11 ± 0.91 | 4.12 ± 0.91 | | |
| **Subjective Norms** | 3.40 ± 0.99 | 3.42 ± 1.0 | 3.33 ± 0.94 | 0.24 | 0.91 |
| Most people whose opinions matter to me think that I should limit my child's intake of sugar-sweetened drinks to a small serving or none each day. | 3.25 ± 1.19 | 3.26 ± 1.20 | 3.21 ± 1.14 | | |
| Most people whose opinions matter to me think that I should limit my child's intake of sweetened foods to a small serving or none each day. | 3.31 ± 1.5 | 3.32 ± 1.16 | 3.27 ± 1.14 | | |
| Most people whose opinions matter to me think that I should limit my child's screen time to less than 2 hours each day. | 3.19 ± 1.13 | 3.22 ± 1.12 | 3.06 ± 1.14 | | |
| Most people whose opinions matter to me think that I should get my child to spend his or her free time each day doing something physically active. | 3.62 ± 1.15 | 3.64 ± 1.15 | 3.53 ± 1.24 | | |
| Most people whose opinions matter to me think that I should have my child get enough sleep each day. | 3.67 ± 1.09 | 3.68 ± 1.10 | 3.62 ± 1.07 | | |
| **Perceived Behavioral Control** | 4.26 ± 0.54 | 4.28 ± 0.53 | 4.17 ± 0.57 | 0.003 | 0.73 |
| I am confident that I can limit my child's intake of sugar-sweetened drinks to a small serving or none each day. | 4.39 ± 0.71 | 4.41 ± 0.70 | 4.30 ± 0.75 |  |  |
| I am confident that I can limit my child's intake of sweetened foods to a small serving or none each day. | 4.27 ± 0.73 | 4.27 ± 0.74 | 4.23 ± 0.70 |  |  |
| I am confident that I can limit my child's screen time to less than 2 hours each day. | 3.94 ± 0.94 | 3.97 ± 0.92 | 3.78 ± 1.02 |  |  |
| I am confident that I can get my child to spend his or her free time each day doing something physically active. | 4.45 ± 0.61 | 4.46 ± 0.60 | 4.38 ± 0.63 |  |  |
| I am confident that I can have my child get enough sleep each day. | 4.26 ± 0.81 | 4.28 ± 0.79 | 4.16 ± 0.91 |  |  |
| **Behavioral Intention** | 4.23 ± 0.56 | 4.25 ± 0.56 | 4.17 ± 0.57 | 0.02 | 0.78 |
| I intend to limit my child's intake of sugar-sweetened drinks to a small serving or none each day. | 4.26 ± 0.81 | 4.27 ± 0.80 | 4.21 ± 0.83 | - | - |
| I intend to limit my child's intake of sweetened foods to a small serving or none each day. | 4.19 ± 0.78 | 4.20 ± 0.78 | 4.14 ± 0.81 | - | - |
| I intend to limit my child's screen time to less than 2 hours each day. | 3.84 ± 0.99 | 3.87 ± 0.97 | 3.73 ± 1.05 | - | - |
| I intend to get my child to spend his or her free time each day doing something physically active. | 4.41 ± 0.61 | 4.43 ± 0.60 | 4.34 ± 0.64 | - | - |
| I intend to have my child get enough sleep each day. | 4.48 ± 0.60 | 4.49 ± 0.59 | 4.42 ± 0.64 | - | - |

<sup>a</sup> Item response options ranged from 1 (strongly disagree) to 5 (strongly agree).
<sup>b</sup> *P* values for t-tests examining differences in TPB constructs by household food security status.

Table 4 presents unadjusted and adjusted logistic regression models for associations of TPB constructs related to parent engagement in health-promoting behaviors to prevent obesity and household food security. In unadjusted logistic regression models, parents who reported greater perceived behavioral control (OR = 1.44, CI: 1.14 -1.82) had significantly higher odds of experiencing household food security. In adjusted logistic regression models controlling for parent education and Medicaid participation, this finding remained significant (OR =1.45, CI: 1.15 - 1.85). Parent-reported attitude and subjective norm were not significantly associated with household food security status.

### Food Resource Management and Food Security

Table 3 presents descriptives for parent food resource management self-confidence and behaviors by household food security status. The mean value of food resource management self-confidence among food secure households (M = 3.97, SD = 0.86) was significantly higher compared to food insecure households (M = 3.35, SD = 0.93; *P* < 0.001).

**Table 3.** Descriptives for parent food resource management self-confidence and behaviors by household food security status (n = 1622).

| Construct and Items<sup>a</sup> | Total Sample<br>Mean ± SD<br>(n = 1622) | Food Secure<br>Mean ± SD<br>(n = 1343) | Food Insecure<br>Mean ± SD<br>(n = 278) | *P*<br>Value<sup>b</sup> | Alpha |
| --- | --- | --- | --- | --- | --- |
| **Self-confidence** | 3.87 ± 0.91 | 3.97 ± 0.86 | 3.35 ± 0.93 | <0.001 | 0.85 |
| How confident are you that you can buy healthy foods for your family on a budget? | 3.89 ± 1.07 | 3.99 ± 1.02 | 3.39 ± 1.18 | - | - |
| How confident are you that you can choose the best priced form of fruits and vegetables? | 3.96 ± 1.01 | 4.03 ± 0.98 | 3.64 ± 1.08 | - | - |
| How confident are you that you can make your food money last all month? | 3.87 ± 1.17 | 4.06 ± 1.08 | 2.98 ± 1.22 | - | - |
| How confident are you that you can make low cost meals? | 3.75 ± 1.10 | 3.82 ± 1.09 | 3.41 ± 1.10 | - | - |
| **Behaviors** | 3.71 ± 0.67 | 3.70 ± 0.69 | 3.73 ± 0.72 | 0.41 | 0.71 |
| How often do you plan meals ahead of time? | 3.49 ± 0.90 | 3.53 ± 0.88 | 3.34 ± 0.99 | - | - |
| How often do you use a grocery list when you go grocery shopping? | 3.89 ± 1.06 | 3.92 ± 1.05 | 3.75 ± 1.06 | - | - |
| How often do you compare prices before you buy food? | 3.55 ± 1.19 | 3.50 ± 1.20 | 3.80 ± 1.12 | - | - |
| How often do you look in the refrigerator or pantry before you go shopping to see what food you need? | 4.25 ± 0.82 | 4.25 ± 0.83 | 4.23 ± 0.82 | - | - |
| How often do you change your grocery list in the store to include foods that are on sale? | 3.32 ± 1.07 | 3.28 ± 1.07 | 3.54 ± 1.06 | - | - |

<sup>a</sup> Response options for Self-confidence options ranged from 1 (not at all confident) to 5 (very confident) and Behaviors ranged from 1 (never) to 5 (always).
<sup>b</sup> *P* values for t-tests examining differences in food resource management constructs by household food security status.

Table 4 presents unadjusted and adjusted logistic regression models for associations of parent food resource management constructs and household food security. In unadjusted logistic regression models, parents who reported greater self-confidence (OR = 2.08, CI: 1.80 - 2.41), but not behaviors, had significantly higher odds of experiencing household food security. In adjusted logistic regression models controlling for parent education and Medicaid participation, this association remained significant (OR = 2.12, CI: 1.83 - 2.46).

**Table 4.** Logistic regression results for Theory of Planned Behavior (TBP) constructs related to parent engagement in health-promoting behaviors to prevent obesity and food resource management constructs and household food security (n = 1622).

| Constructs | Unadjusted Logistic Regression |  | Adjusted Logistic Regression<sup>a</sup> |  |
| --- | --- | --- | --- | --- |
|  | OR | 95% CI | OR | 95% CI |
| Theory of Planned Behavior |  |  |  |  |
| Attitude | 1.04 | 0.84 - 1.27 | - | - |
| Subjective norm | 1.10 | 0.96 - 1.25 | - | - |
| Perceived behavioral control | 1.44<sup>b</sup> | 1.14 - 1.82 | 1.45<sup>b</sup> | 1.15 - 1.85 |
| Food resource management |  |  |  |  |
| Self-confidence | 2.08<sup>b</sup> | 1.80 - 2.41 | 2.12<sup>b</sup> | 1.83 - 2.46 |
| Behaviors | 0.93 | 0.77 - 1.12 | - | - |

<sup>a</sup> Models adjusted for parent education and Medicaid participation.
<sup>b</sup> Model statistically significant when confidence interval do not overlap one.
Note: OR, odds ratio; CI, confidence interval

## DISCUSSION

This study examined associations between parent engagement in health-promoting behaviors and food resource management with household food security status among parents with young children living in rural, lower-income contexts. Our findings show that a greater proportion of parents in this study experienced household food insecurity during the past year when compared to the national average and rural households alone.^1^ Our findings showed that greater perceived behavioral control, a TPB construct, and food resource management self-confidence were associated with higher odds of household food security in adjusted models. Understanding which parent attitudes and behaviors are associated with household food security can be used to inform health promotion efforts that target nutrition, health, and well-being among families living in lower-income contexts.

The TPB, a widely used theory to understand health behaviors^18^, has been used to explore parent behaviors such as food purchasing^21^, feeding decisions^22^, and obesity prevention^23^ for their young children. To our knowledge, the current study is the first to examine associations between TPB constructs related to parent engagement in health-promoting behaviors and household food security status. In the current study, parent reports of perceived behavioral control and behavioral intention were higher in food secure households compared to food insecure households. Parent perceived behavioral control was also associated with higher odds of experiencing household food security. Thus, parents in this sample of rural, lower-income families who were more confident engaging in health-promoting behaviors around nutrition, physical activity, sleep, and screen time among their young children were more likely to experience household food security. In addition, a study that included mothers of preschool-aged children showed that perceived behavioral control was associated with higher child fruit and vegetable intake and lower unhealthy snack food intake.^35^ Given that perceived behavioral control is a predictor of behavioral intention and subsequent engagement in a behavior, according to the TPB, ^18^ this finding is important because greater perceived behavioral control may help people persevere during times of hardship.

Findings from previous studies have shown associations between other TPB constructs and parent health-promoting behaviors. A study by Capasso et al., extended the TPB to include mother’s information seeking behavior (i.e., reading food labels) to inform food purchases for their children and found that mother’s attitude, but not perceived behavioral control, predicted behavioral intention.^21^ Our findings showed that attitude and subjective norms were not significantly associated with household food security. The lack of variation in attitude and subjective norms toward health-promoting behaviors, which were high in both households that experienced food security and those that did not, may partially explain this finding. In addition, subjective norms may not be as salient for parents who want to support their child’s growth and development but may experience barriers and stressors related to economic hardship.^17^ A recent study by van der Velde et al. extended the TPB to include financial scarcity and food insecurity to understand diet quality among adults showed that including these factors better explain diet quality.^36^ Future studies that use the TPB to understand parent engagement in health-promoting behaviors for their young children should include financial scarcity and food security status to extend the TPB to understand structural relationships among these constructs.

Food resource management behaviors such as meal planning, selecting nutritious foods, and stretching limited food budgets may help improve household food and nutrition security.^27^ Nutrition education programs designed to improve food resource management self-confidence have been associated with lower odds of household food insecurity.^33,37^ In line with this research, our findings showed that parent food resource management self-confidence was associated with household food security. Parents in this sample who experienced household food security, reported higher self-confidence related to purchasing nutritious foods on a budget, preparing low cost meals, and making food money last all month. In contrast, our findings showed that food resource management behaviors were not significantly associated with household food security.^37^ However, a study exploring the impact of *Cooking Matters for Adults*, a nutrition education program for adults living in low-income contexts, showed lasting improvements in food resource management behaviors that led to less concern that food might run out before participants had the resources to buy more.^26^ Although these behaviors were not associated with food security in our sample, food resource management behaviors have been associated with higher diet quality^38,39^, thus helping parents develop and engage in these behaviors may help improve health outcomes. In addition, in our increasingly complex and obesogenic food enviroment^40^, helping parents develop and engage in behaviors such as meal planning and purchasing nutritious foods on a budget may also increase self-confidence and promote the health and well-being of their families.

Although this study has many strengths, including examining factors associated with household food security status in a large sample of parents of young children living in rural lower income contexts who are at risk for poor health outcomes, it is not without limitations. First, baseline data for the ENCIRCLE study were collected between 2020-2023 during the COVID-19 pandemic and it is unknown how pandemic-related relief programs impacted food security status in this sample. Second, the TPB measure was adapted for a lower literacy audience, and although the adapted measure was reviewed by members of the research team and community advisory board, it was not validated. However, the adapted measure had good to acceptable reliability. Third, this study used the 6-item short-form US Household Food Security Survey Module,^34^ which accesses household-level food security status, thus child food security status is unknown. Lastly, the current study was cross-sectional, thus causality cannot be determined when examining associations between parent health-promoting behaviors and food resource management with household food security. In addition, the impact of unmeasured confounding by factors such as parent age and neighborhood food access are unknown.

### Implications for Research and Practice

Improving food and nutrition security has received much attention. Efforts to increase funding for and access to federal nutrition assistance programs are underway, although policy, systems, and environmental changes take time to implement. In the meantime, health promotion efforts that aim to improve parents perceived behavioral control and food resource management self-efficacy may help promote food security and safeguard rural, lower-income families. Improving these psychosocial factors may help parents engage in health-promoting behaviors such as providing healthy foods and stretching limited resources during times of economic hardship.

## Acknowledgments

All authors would like to thank the courageous parents with young children who participated in this study and the dedicated researchers who were responsible for data collection and curation. This would not have been possible without each of you, thank you.

## Author Contributions

Conceptualization, AMM, LBD; Methodology, AMM, LBD; Analysis, AMM; Writing – Original Draft Preparation, AMM; Writing – Intellectual Content, Review & Editing, AMM, JSS, MNP, JFH, CM, LBD; Funding Acquisition, LBD (PI of the main study).

## Institutional Review Board Statement

The study was conducted according to the guidelines of the Declaration of Helsinki and approved by the Institutional Review Board of Geisinger.

## Informed Consent Statement

Informed consent was obtained from all subjects involved in the study.

## Data Availability Statement

De-identified data are available upon reasonable request and with a data sharing agreement in place from AMM.

